# Dimensionality and homogeneity of the Colombian version of the Cocaine Craving Questionnaire (CCQ-N-10)

**DOI:** 10.1101/2021.12.07.21267420

**Authors:** Carlos Arturo Cassiani-Miranda, Orlando Scoppetta, Tito Cesar Quintero-Gómez, Eduard Arraut-Camargo, Guillermo Castaño-Pérez, Diego Fernando Cabanzo-Arenas, Adalberto Campo-Arias

**Affiliations:** Universidad de Santander, Bucaramanga, Colombia; Corporación Nuevos Rumbos. Bogotá, Colombia; Universidad Industrial de Santander, Bucaramanga, Colombia; Hospital Psiquiátrico San Camilo, Bucaramanga-Colombia; Universidad CES, Medellín, Colombia; Universidad del Magdalena, Santa Marta, Colombia

**Keywords:** Psychometrics, craving, cocaine, Colombia

## Abstract

**Background and aim:** It is essential to have a validated instrument to measure craving for cocaine use. However, in Colombia, there is no instrument to screen cocaine craving. The study aimed to determine the homogeneity and dimensionality of the CCQ-N-10 in patients with cocaine use disorder in Colombia.

**Materials and methods:** An adaptation and subsequent analysis of the psychometric properties of the CCQ-N-10 scale was carried out with 102 hospitalized or outpatient adults diagnosed with substance use disorders in addiction units in Bucaramanga, Colombia. Internal consistency and construct validity were estimated by confirmatory factor analysis and correlation analysis with scales with similar objectives.

**Results:** An omega coefficient of 0.93 was obtained, and adjustment indicators of the confirmatory model were acceptable (RMSEA of 0.08, CFI and TLI of 0.99) when two of the original scale items were removed from the original scale analysis.

**Conclusions:** This study shows that the craving scale reduced to eight items can be helpful to assess the construct in the Colombian population; However, the small sample size makes it challenging to carry out other analyzes to corroborate its psychometric properties.

## 1. Introduction

Craving is a central aspect of cocaine use disorders (CUD) and is strongly associated with relapses [1]. Consequently, valid and reliable measurement of craving is essential for CUD research and practice [2]. Evaluation of the efficacy of CUD interventions should benefit from an adequate assessment of craving [3].

There are several self-administered scales, among which are the Voris Cocaine Craving Scale, VCCS [4], the Cocaine Craving Questionnaire, CCQ [5], or the Cocaine Craving Scale, CCS [6]. The CCQ is the most used for validity and solid theoretical support [2]. However, it has 45 items, limiting its use in the clinical setting. Abbreviated versions of the CCQ have been evaluated [1, 7, 8], such as the 10-item version (CCQ-N-10) [7] from the original version of the CCQ-Now [5]. The CCQ-N-10 is a self-report instrument with a seven-point visual analog scale response pattern as strongly disagree” on one pole and “strongly agree” on the other, obtaining a score of 0 to 6 points according to the degree of agreement or disagreement in each statement [7]. The CCQ-N-10 has shown adequate convergent validity with measures of craving such as VCCS (r of 0.47) and adequate predictive validity [1]. CCQ-N-10 was adapted and validated in Spanish [9, 10], showing adequate internal consistency (Cronbach’s alpha between 0.87 and 0.95). Moreover, CCQ-N-10 presented a stable one-dimensional structure (CFI of 0.91 and SRMR of 0.06), adequate test-retest validity (Intraclass correlation coefficient of 0.59, p < 0.001), and convergent validity with other related constructs (CCQ-CCS r of 0.64, CCQ-EVA r of 0.65; CCQ-SDS r of 0.53, and CCQ-ICP r of 0.50).

It is necessary to have reliable and brief instruments for assessing cocaine craving in the clinical setting. There are no published data on the validity or reliability of any instrument used to evaluate cocaine craving in Colombia.

This research aimed to determine the homogeneity and dimensionality of the CCQ-N-10 in patients with CUD in Colombia.

## 2. Materials and methods

### 2.1. Design

Validation study of a scale without reference criteria.

### 2.2. Population and sample

One hundred and two hospitalized or outpatient adults diagnosed with CBT were recruited in addiction units in Bucaramanga, Colombia. The mean age was 29 years (deviation of 9.4), 83% were men, 90.2% came from urban areas, 81.2 belonged to socioeconomic strata 1 to 3, only 3% had 11 years of schooling, 77.5% were single, and 11.8% married.

### 2.3. Measurements

#### 2.3.1 Cocaine craving questionnaire-short current version (CCQ-N-10)

It was developed by Sussner et al. [7] from the original version of the 45-item CCQ-Now [5], retaining the ten items with the highest factorial weight in the first factor (“desire to consume”). CCQ-N-10 has a response format in the form of a seven-point visual analog scale depending on the degree of agreement or disagreement with each statement. Scores for each item range from a minimum of 0 to a maximum of 6 points. The items raised inversely will be recorded, reversing the order of the indicated value. The recoded items were 4 and 7: “I think I could resist and not use cocaine now.” and “I do not want to use cocaine right now.” The version was adapted by Muñoz et al. [8] with the authors’ permission.

#### 2.3.2 Cocaine Craving Scale (CCS)

The CCS was developed by Weiss et al. [6], and it is composed of 5 items with a response scale of 0 to 9 points. It is one-dimensional and has good internal consistency (alpha between 0.82 and 0.94). The Spanish adaptation made by Tejero et al. [11].

#### 2.3.3 Brief Spanish Cocaine Craving Questionnaire-General (BCCQ-G)

Developed from CCQ-G that investigates the level of craving the previous week in cocaine users through 45 items scored in a Likert-type response format from 1 to 7 that measure the degree agree or disagree that the interviewee has about each statement [5]. This scale is adapted with 12 items in the Spanish population (n=183). The BCCQ-G showed excellent fit indicators in the CFA (χ2=57.48, p=0.16, SRMSR=0.06, CFI=0.98.) and good internal consistency (Cronbach’s alpha of 0.89). The 12-item version adapted in Spain was used with the authors’ permission [8].

### 2.4. Procedure

The project was approved by the ethics committee and the research department of the Universidad de Santander. The authors used the version of CCQ-N-10 validated by Castillo et al. [10], from which experts carried out the validity of appearance and content following international recommendations [12]: six experts in addictions and research (four psychiatrists and one psychologist) were consulted to evaluate the relevance and pertinence of the items. Open questions were asked about the usefulness of the scale to detect craving in cocaine users in this population, and language corrections were requested to the instrument questions. According to the experts’ recommendations, grammar adjustments were made through the research group’s discussion group. The scale adapted for Colombia was evaluated in a pilot test with ten patients by two different evaluators. The patients evaluated were people who attended outpatient or intramural treatment for addictive disorders due to problematic cocaine use, and the evaluators were psychiatrists or psychologists. In this phase, aspects related to the particularities of the items, the application time, and the scale’s usefulness were analyzed [13]. There were no significant problems in understanding, so no additional adjustments were made to the scale.

### 2.5. Analysis of data

According to previous studies, a confirmatory factor analysis (CFA) was performed with structural covariance techniques to verify the unidimensionality of the CCQ-N-10 [7]. The degree of adjustment to the sample data of the hypothesized theoretical model was estimated by estimating the parameters according to the maximum likelihood method. The goodness of fit of the model was evaluated with the following indicators: 1) goodness of fit index (GFI), 2) Bentler’s comparative fit index (CFI), 3) root mean square error of approximation (RMSEA). For a parsimonious adjustment, the GFI and CFI values should be close to 0.90, and the RMSEA should be around 0.08 [14].

Cronbach’s alpha [15] and McDonald’s omega [16] were calculated to estimate the internal consistency of the global scale and with the omission of each item. Convergent validity was evaluated by comparing the CCQ-N-10 scores with other scales to measure craving for cocaine (BCCQ-G, CCS) using the Spearman or Pearson correlation according to the data distribution. The analysis was done with R statistical program [17] and MPLUS, version 7 [18].

## 3. Results

### 3.1. Internal consistency

Cronbach’s alpha and McDonald’s omega were 0.93. The item-rest of items correlation ranged from 0.66 to 0.87 (Table 1).

**Table 1.**
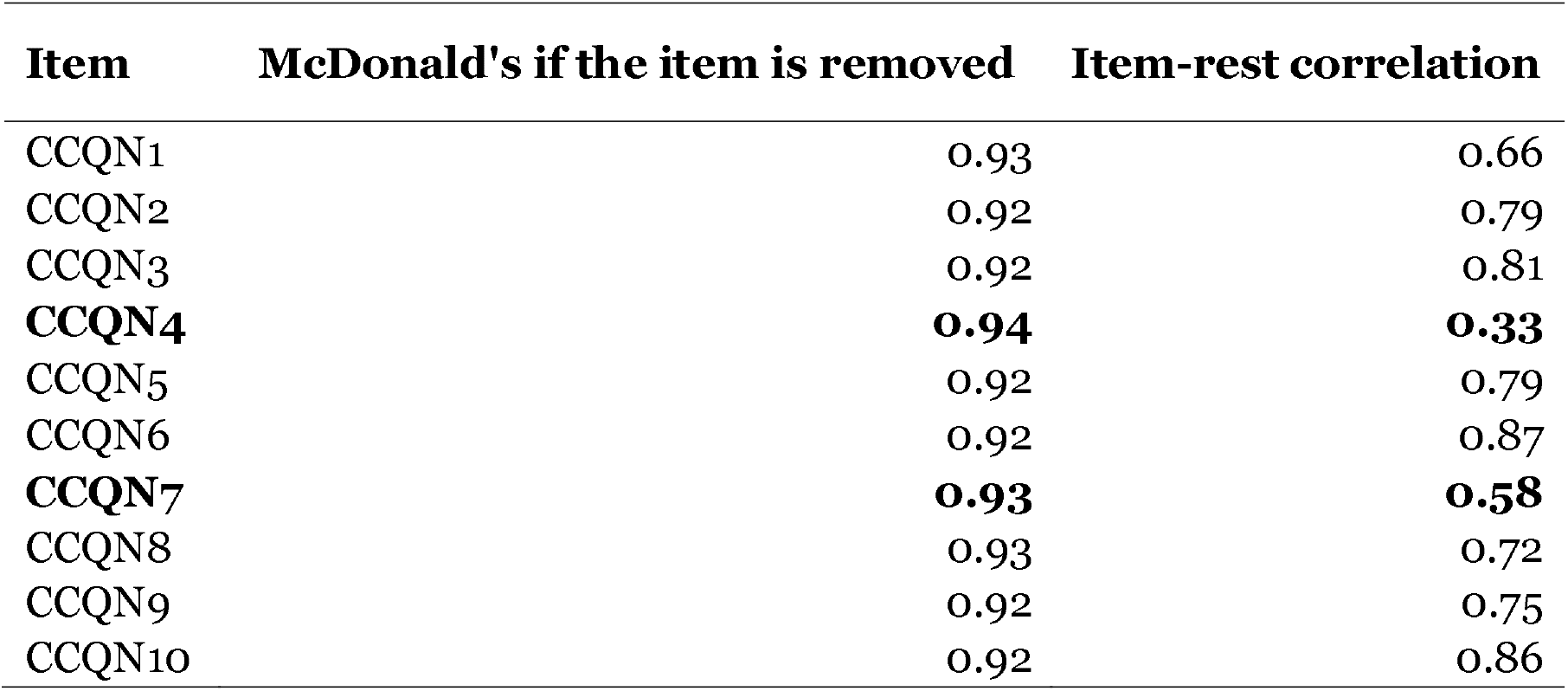
Item and item-item correlations of the CCQ-N-8.

| Item | McDonald's if the item is removed | Item-rest correlation |
| --- | --- | --- |
| CCQN1 | 0.93 | 0.66 |
| CCQN2 | 0.92 | 0.79 |
| CCQN3 | 0.92 | 0.81 |
| **CCQN4** | **0.94** | **0.33** |
| CCQN5 | 0.92 | 0.79 |
| CCQN6 | 0.92 | 0.87 |
| **CCQN7** | **0.93** | **0.58** |
| CCQN8 | 0.93 | 0.72 |
| CCQN9 | 0.92 | 0.75 |
| CCQN10 | 0.92 | 0.86 |

### 3.2. Factor analyses

An AFC was made with various models. The model was adjusted without items 4 and 7. In table 1, it was observed that these items had the lowest correlations to the rest of the instrument.

Table 2 shows the standardized loads and the square correlations of each item. Standardized regression coefficients show how much each item changes in standard deviations by changing one standard deviation of the factor. Square correlations indicate the percentage of the variance explained for the item.

**Table 2.** Standardised loadings and squared correlations for each item of the CCQ-N-8.

| **Item** | **Standardised loads** | **Squared correlations** |
| --- | --- | --- |
| CCQN1 | 0.71 | 0.51 |
| CCQN2 | 0.84 | 0.70 |
| CCQN3 | 0.93 | 0.86 |
| CCQN5 | 0.93 | 0.86 |
| CCQN6 | 0.95 | 0.91 |
| CCQN8 | 0.89 | 0.79 |
| CCQN9 | 0.88 | 0.78 |
| CCQN10 | 0.95 | 0.90 |

All p values <0.01.

The fit indicators of the one-dimensional model of the proposed version (CCQ-N-8) were: RMSEA of 0.08 (90CI% 0.01-0.12), CFI, and TLI of 0.99. The original version (CCQ-N-10) adjustment indicators were: RMSEA of 0.18 (90%CI 0.15-0.21), CFI of 0.96, and TLI of 0.95.

### 3.3. Convergent validity

The Spearman correlation coefficients of CCQ-N-8 were 0.72 with the CCS and 0.59 with the BCCQ-G.

## 4. Discussion

This study shows that CCQ-N-8 presents better goodness of fit indicators than ten items (CCQ-N-10); it showed a stable one-dimensional structure, high internal consistency, and adequate convergent validity.

Cronbach’s alpha and MacDonald’s omega for CCQ-N-8 were 0.93, indicating high internal consistency. Although these two coefficients are comparable, McDonald’s omega is the best estimator of internal consistency when the principle of tau equivalence is violated [19], as is often the case on scales such as CCQ-N-10 [7]. Although the validation studies of CCQ-N-10 have not reported omega, the internal consistency of CCQ-N-8 is comparable to that found for CCQ-N-10 [7, 9, 10].

An excellent measuring instrument must show acceptable dimensionality and homogeneity to ensure maximum measurement precision [20]. The CFA showed low indicators of goodness of fit for CCQ-N-10, so several scale models were tested, obtaining the most parsimonious fit with the 8-item version (without items 4 and 7), which showed high correlations, lower with the full scale (0.33 and 0.58 respectively) compared to the others whose correlations were between 0.66 and 0.87. When comparing the goodness of fit indicators for a version of 8 and 10 items, the CCQ-N-8 showed more appropriate goodness of fit indicators (RMSEA of 0.08, CFI and TLI of 0.99 vs. RMSEA of 0.18, CFI of 0.96, and TLI of 0.95). These items require recoding, which may affect the scale’s performance because items with negation in the statement or require recoding tend to affect the individual’s ability to answer adequately, compromising validity and reliability [21].

The CCQ-N-8 showed a strong positive correlation with CCS (r of 0.72) and moderated with BCCQ-G (r of 0.59), indicating that CCN-N-8 has adequate convergent validity. Even the correlations with CCS were higher than that observed with the original scale [7].

### 4.1. Strengths and limitations of the study

This study presents a new version of the instrument used to evaluate craving for the first time in a Latin American population. However, it has limitations: The small sample size, since to obtain adequate precision and power in estimating the fit indices of the model, 250 to 500 participants are required [22]. The instrument’s stability [test-retest reliability] was not evaluated; this information is vital for patient follow-up when performing repeated evaluations.

The differential functioning of the items and the construct validity were not evaluated through models based on the item response theory that provide additional elements for the refinement of the scales [23]. However, having shorter versions of measurement instruments can offer some advantages in research and the clinical setting: blunt instruments are usually one-dimensional and show more reproducible psychometric indicators in different contexts [24]. In addition, short scales are beneficial for clinical practice settings, where time and resources are limited, and for research studies where the burden of assessment needs to be minimized [25].

### 4.2. Practical issues

Finally, it is necessary to remember that the performance of the instruments can vary significantly between samples or populations, which can threaten the validity and reliability of the measurement and implies the need for continuous evaluation and improvement of the instruments [20].

### 4.3. Conclusions

The 8-item version of the current cocaine craving questionnaire is a one-dimensional instrument with high internal consistency and convergent validity with other validated scales to measure cocaine craving. CCQ-N-8 is recommended to evaluate the Colombian population for cocaine craving. It is necessary to corroborate its factorial structure with a more appropriate sample size in Spanish-speaking countries and other languages.

## Data Availability

All data produced in the present study are available upon reasonable request to the corresponding author.

